# Quality of Chronic Disease–Related Health Videos Across Social Media Platforms: A Systematic Review and Meta-analysis

**DOI:** 10.64898/2026.07.14.26358040

**Authors:** Ruofan Liu, Yuxiang Xu, Maomao Zhang, Yunfei Li, Xinyang Wang, Chuanbing Huang

**Affiliations:** The First Affiliated Hospital of Anhui University of Traditional Chinese Medicine, Hefei, China; School of Chinese Medicine, Bozhou University, Bozhou, China; Key Laboratory of Xin’an Medicine, Ministry of Education, Anhui University of Chinese Medicine, Hefei, China

**Author notes:** **Corresponding authors:** Chuanbing Huang, PhD, Address: The First Affiliated Hospital of Anhui University of Traditional Chinese Medicine, Hefei, Anhui, 230000, China.

## Abstract

**Background:** Social media videos have become one of the major sources of health information for individuals living with chronic diseases. Although numerous cross-sectional studies have evaluated the quality of health-related videos across different platforms, the overall quality of chronic disease–related videos and the determinants underlying quality variation remain unclear.

**Objective:** To systematically evaluate the quality of chronic disease–related health videos across major global and Chinese social media platforms and to identify potential determinants of video quality using multivariable meta-regression.

**Methods:** This systematic review and meta-analysis searched PubMed, Embase, and Web of Science from database inception to April 30, 2026, for cross-sectional studies evaluating Chinese- and English-language health videos. Scores from the DISCERN instrument, the Global Quality Scale (GQS), and the Journal of the American Medical Association (JAMA) benchmark criteria were standardized to a 0–100 scale and quantitatively synthesized using random-effects models. Prespecified subgroup analyses and multivariable meta-regression were conducted to explore potential sources of heterogeneity, including platform region, platform type, disease category, video duration, professional background of content creators, and audience engagement.

**Results:** A total of 88 studies involving 18,688 videos were included. Overall methodological quality was suboptimal, with a pooled standardized DISCERN score of 50.61 (95% CI, 48.04–53.18), accompanied by substantial between-study heterogeneity (I² = 99.3%). Videos hosted on international platforms achieved significantly higher quality scores than those on Chinese platforms (54.68 vs. 48.21; *P* = 0.008). Multivariable meta-regression demonstrated that conventional predictors—including video duration, the proportion of physician creators, and audience engagement—were not independently associated with video quality (*P* > 0.05). Importantly, the final model explained only 9.06% of the between-study heterogeneity (R² = 9.06%), indicating that conventional content- and creator-level characteristics account for only a small proportion of the observed variability in video quality.

**Conclusions:** Traditional predictors, including creator professionalism, video duration, disease category, and audience engagement, have limited ability to explain variation in the quality of online health videos. Although platform region emerged as the only significant moderator, the multivariable model explained only a small fraction of the observed heterogeneity, suggesting that the principal determinants of health information quality remain largely unexplained.

## 1. Introduction

Chronic conditions requiring long-term management have become one of the greatest threats to global public health and socioeconomic sustainability. Epidemiological evidence indicates that chronic diseases account for more than 74% of all deaths worldwide, and this proportion approaches 90% in countries with rapidly aging populations(1). Beyond their direct contribution to mortality, chronic diseases are characterized by prolonged disease courses and limited curative options, resulting in substantial disability-adjusted life years (DALYs) and enormous economic burdens.

Moreover, this burden is distributed unequally across socioeconomic groups. For patients living in low-income settings or regions with limited healthcare resources, geographical barriers, inadequate access to high-quality medical services, and the high risk of catastrophic health expenditure (CHE) substantially restrict opportunities to obtain professional medical guidance through conventional healthcare systems(2).

Given the persistent burden of chronic diseases, effective self-management and continuous health education outside clinical settings have become essential components of disease control, while also reducing pressure on healthcare systems and household financial burdens (3). The widespread adoption of smartphones and online video platforms has provided patients with an inexpensive and highly accessible alternative source of health information (4). For many individuals living with chronic diseases, social media has evolved beyond an entertainment medium to become one of the primary channels for obtaining disease management knowledge and health-related support (5).

Advances in mobile applications, artificial intelligence (AI), and online video technologies have driven the rapid expansion of digital health communication in recent years (6). However, the scientific accuracy and reliability of health information disseminated through these channels remain highly variable, leading to increasing concern regarding information quality (7). Online videos, as one of the most influential forms of health communication, present both opportunities and challenges. On one hand, they have the potential to overcome geographical barriers and reduce disparities in access to health information. On the other hand, the absence of rigorous peer review and the algorithm-driven nature of online platforms raise substantial concerns regarding the credibility of the information they disseminate (8).

More importantly, recommendation algorithms are inherently designed to maximize user engagement and traffic, often prioritizing popularity over scientific rigor or evidence-based accuracy. Compared with individuals affected by acute illnesses, patients with chronic diseases—who require long-term disease management and often face sustained financial burdens—are particularly vulnerable to low-quality health information (9). Reliance on inaccurate or misleading medical advice may delay appropriate treatment, worsen disease outcomes, and increase unnecessary healthcare expenditures, thereby exacerbating the risk of medical impoverishment.

Given the heavy reliance of individuals with chronic diseases on online health videos and their heightened vulnerability to misinformation, evaluating the dissemination of health information alone is no longer sufficient; systematic assessment of the quality of medical information available on social media has become increasingly important. Although studies evaluating the quality of health-related videos on major online platforms have grown rapidly, most have focused on a single disease or a single platform using cross-sectional designs (10, 11). Consequently, comprehensive evidence synthesizing video quality across multiple diseases and platforms remains lacking.

Therefore, this study aimed to comprehensively evaluate the quality and reliability of chronic disease–related health videos across major social media platforms using rigorous evidence synthesis methods. Our findings are expected to provide a clearer understanding of the current landscape of digital health communication and to generate evidence that may inform public health policy, guide healthcare professionals in producing evidence-based educational content, and protect vulnerable patient populations from health misinformation.

## 2. Methods

### 2.1 Study Design and Registration

This systematic review and meta-analysis was conducted and reported in accordance with the 2020 Preferred Reporting Items for Systematic Reviews and Meta-Analyses (PRISMA 2020) statement. The study protocol was prospectively registered in the International Prospective Register of Systematic Reviews (PROSPERO) (Registration No. CRD420261345384).

### 2.2 Search Strategy and Eligibility Criteria

A comprehensive literature search was conducted in PubMed, Web of Science, Embase, and the Cochrane Library from database inception to April 30, 2026, to identify studies evaluating the quality of online videos related to chronic diseases. Search strategies combined controlled vocabulary and free-text terms, including keywords related to social media videos, health information, quality, DISCERN, Global Quality Scale (GQS), and the JAMA benchmark criteria. Because chronic diseases encompass a broad spectrum of heterogeneous conditions, a platform-oriented search strategy was adopted rather than disease-specific searches to maximize sensitivity. Eligible studies were subsequently identified through predefined inclusion and exclusion criteria. Only peer-reviewed original studies published in English were included. Eligible studies evaluated health-related videos on major Chinese platforms (Bilibili, Douyin, Rednote, and Kwai) or international platforms (YouTube and TikTok). The complete search strategy is presented in Supplementary Appendix 1 Full Search Strategies.

Given that platform popularity is closely related to the dissemination and influence of health information, six major video platforms (Bilibili, Douyin, Rednote, Kwai, YouTube, and TikTok) were prespecified. Studies were considered eligible if they evaluated chronic disease–related health videos on one or more of these platforms and reported standardized video characteristics and/or validated measures of information quality.

All retrieved records were imported into EndNote 21 for duplicate removal. Two reviewers (Liu and Xu) independently screened studies by title and abstract, followed by full-text assessment. Disagreements were resolved through discussion with a third reviewer (Li).

Eligibility criteria were established according to the PICO framework. Chronic diseases were defined according to the World Health Organization (WHO) and the Centers for Disease Control and Prevention (CDC) as conditions lasting at least one year that require ongoing medical care and/or limit activities of daily living. Specifically, we included diseases requiring long-term clinical monitoring, continuous pharmacological treatment, or sustained patient self-management, such as cardiovascular diseases, metabolic disorders, and chronic musculoskeletal diseases.

Studies were excluded if they focused on (1) acute or life-threatening medical emergencies, (2) self-limiting or acute infectious diseases, or (3) short-term surgical interventions. Case reports, editorials, letters, expert opinions, reviews, animal studies, and studies lacking full-text articles or extractable data were also excluded.

### 2.3 Data Extraction and Data Processing

Data extracted from each eligible study included author, country, publication year, platform, disease category, sample size, search date, video duration, number of likes, comments, favorites, shares, proportion of professional creators, and quality assessment outcomes (DISCERN, GQS, and JAMA). To ensure statistical compatibility, only studies reporting continuous variables as mean (SD) or median (IQR) were included. For studies evaluating multiple platforms, outcome data were extracted separately for each platform whenever available. Data extraction was performed independently by two reviewers (Liu and Xu), with any discrepancies resolved through consensus.

To harmonize different reporting formats and quality assessment instruments, rigorous data processing procedures were performed. For studies reporting continuous outcomes as median (IQR), sample means and standard deviations were estimated using the validated conversion method proposed by Wan et al.(12), enabling inclusion in quantitative synthesis. Because different quality assessment instruments employ different scoring ranges (e.g., DISCERN, GQS, and JAMA), standardized mean scores were generated by proportionally rescaling all scores to a unified 0–100 scale, thereby facilitating direct comparisons across studies.

Specifically, each reported mean score and its corresponding standard deviation were divided by the maximum possible score of the respective instrument and multiplied by 100. This proportional transformation preserves the relative distribution of scores while improving comparability and clinical interpretability across different assessment tools.

For descriptive purposes, standardized scores were categorized into three predefined levels: poor quality (<60 points), acceptable quality (60–79 points), and high quality (≥80 points). These thresholds were established by the investigators to facilitate interpretation and do not represent official cutoffs of the original instruments.

Studies included in quantitative meta-analysis were required to report measures of variance (e.g., standard deviation or 95% confidence interval). Three studies reported an interquartile range of zero, preventing reliable estimation of standard deviations using the Wan method. To avoid assigning these studies disproportionately large statistical weights due to artificially small variances, they were retained in the qualitative synthesis but excluded from quantitative meta-analysis and meta-regression.

### 2.4 Risk of Bias Assessment

Potential publication bias and small-study effects were initially assessed through visual inspection of funnel plot symmetry and subsequently evaluated using Egger’s linear regression test, with P < 0.05 indicating statistically significant asymmetry.

Because most included studies were observational cross-sectional investigations evaluating online video content, methodological quality was assessed using the Joanna Briggs Institute (JBI) Critical Appraisal Checklist for Analytical Cross-Sectional Studies. This instrument evaluates key methodological domains, including sample representativeness, participant description, identification of confounding factors, and validity of outcome measurement.

Two reviewers independently assessed each study as Yes, No, Unclear, or Not applicable for each checklist item. Overall risk of bias was classified as low, moderate, or high according to the proportion of core methodological criteria fulfilled. Inter-reviewer agreement was quantified using Cohen’s kappa coefficient, and discrepancies were resolved through consensus or consultation with a third reviewer (Li).

### 2.5 Data Synthesis and Statistical Analysis

All statistical analyses were performed using R software (version 4.5.1) with the meta and metafor packages. Given the anticipated clinical and methodological heterogeneity arising from differences in platform algorithms, creator characteristics, and disease categories, random-effects models using restricted maximum likelihood (REML) estimation were prespecified for all meta-analyses.

Between-study heterogeneity was assessed using Cochran’s Q test and the I² statistic, with P < 0.10 or I² > 50% considered indicative of substantial heterogeneity. To further characterize the expected range of effect sizes across real-world settings, 95% prediction intervals (PIs) were routinely calculated and reported.

Prespecified subgroup analyses were conducted to explore potential sources of heterogeneity according to (1) platform region (Chinese vs. international platforms), (2) platform type (short-video platforms such as TikTok, Douyin, and Kwai versus longer-form knowledge-sharing platforms such as YouTube and Bilibili), and (3) disease category (e.g., musculoskeletal disorders and endocrine/metabolic diseases).

To further control for potential confounding, multivariable meta-regression models were constructed. Particular emphasis was placed on the proportion of physician creators and average video duration as primary covariates, allowing independent evaluation of the association between professional creator involvement, video length, and overall health information quality.

Leave-one-out sensitivity analyses were conducted by sequentially excluding each individual study to assess the robustness of the pooled estimates and identify potential influential outliers.

Given that all included primary studies were observational cross-sectional designs, the certainty of evidence was not formally assessed using the GRADE approach, which is primarily designed for interventional trials.

## 3 Results

### 3.1 Characteristics of Included Studies

A total of 4,865 records were identified through the database search. After title/abstract screening and full-text assessment, 88 cross-sectional studies met the eligibility criteria and were included in the systematic review and meta-analysis (Figure 1).

Inter-reviewer agreement for both title/abstract screening and full-text assessment was excellent (Cohen’s κ ≥ 0.75). The included studies evaluated 18,688 health-related videos, with a median sample size of 183.5 videos per study. All studies were published between 2021 and 2026, reflecting the recent emergence of this research field.

Most studies originated from China (n = 72, 81.8%), whereas relatively few were conducted in Turkey (n = 11, 12.5%), the United States (n = 4, 4.5%), and India (n = 1, 1.1%) (Figure 3).

Regarding platform distribution, studies predominantly evaluated Chinese domestic platforms (n = 65, 73.9%), whereas investigations of global platforms accounted for only 23 studies (26.1%).

The characteristics of the included studies are summarized in Table 1, and the results of the JBI risk-of-bias assessment are presented in Figure 2.

### 3.2 Meta-analysis Results

#### 3.2.1 Overall Video Quality

Using a random-effects model, 56 studies involving 10,767 videos were included in the pooled analysis of standardized DISCERN scores.

The pooled standardized DISCERN score was 50.61 (95% CI: 48.04–53.18), indicating an overall unsatisfactory level of health information quality. Between-study heterogeneity was extremely high (I² = 99.3%, τ² = 93.40, P < 0.001). The corresponding 95% prediction interval (PI) ranged from 31.06 to 70.15, suggesting substantial variability in video quality across future studies.

For the Global Quality Scale (GQS), 48 studies comprising 10,680 videos were included. The pooled standardized score was 56.93 (95% CI: 54.44–59.43), with similarly substantial heterogeneity (I² = 98.2%, τ² = 74.33, P < 0.001) and a prediction interval of 39.40–74.47.

For the JAMA benchmark criteria, 25 studies involving 6,826 videos were analyzed. The pooled standardized score was 51.85 (95% CI: 47.19–56.52), with extreme heterogeneity (I² = 98.7%, τ² = 138.21, P < 0.001) and a prediction interval of 27.10–76.61.

Overall, all three quality assessment instruments consistently indicated that the methodological quality of online chronic disease videos remained suboptimal. The wide prediction intervals further suggest considerable variability across platforms and studies. Given the substantial heterogeneity, predefined subgroup analyses and meta-regression were subsequently performed to investigate potential sources of between-study variation (Table 2).

#### 3.2.2 Subgroup Analyses

To explore the substantial between-study heterogeneity, predefined subgroup analyses were conducted according to platform region, platform type, and disease category.

##### Platform Region

After controlling for platform type, significant regional differences were observed.

Videos hosted on global platforms demonstrated significantly higher standardized DISCERN scores than those on Chinese domestic platforms (54.68 vs. 48.21; P = 0.008), indicating that platform region was one of the few measurable factors associated with higher information quality (Table 3).

##### Platform-specific Analysis

Health video quality was further compared across the six included platforms.

Most Chinese platforms demonstrated pooled DISCERN scores ranging between 45 and 50, indicating consistently poor information quality. In contrast, greater variation was observed among international platforms. YouTube achieved the highest pooled score (56.91), whereas TikTok demonstrated the lowest (41.22).

However, only three studies evaluated TikTok, and therefore this finding should be interpreted cautiously.

Notably, despite obtaining the highest pooled score, YouTube still failed to reach the predefined threshold for acceptable quality (>60) established in this study.

##### Disease-specific Analysis

To evaluate whether disease category influenced information quality, studies were initially classified into eight disease systems: oncology, cardiovascular diseases, gastrointestinal diseases, endocrine and metabolic diseases, renal and urinary diseases, musculoskeletal diseases, neurological and psychiatric diseases, ophthalmology.

Because of insufficient sample sizes and incomplete data, some diseases were excluded from quantitative subgroup analyses.

The remaining analyses included oncology, musculoskeletal diseases, and ophthalmology, each further stratified according to platform region. Across all disease categories, pooled DISCERN scores remained within the predefined low-quality range (Table 4).

To further examine disease-related effects, diseases were additionally categorized as high-vulnerability (e.g., cancer and chronic musculoskeletal pain) or low-vulnerability (e.g., cardiovascular and ophthalmologic diseases). The pooled DISCERN scores were nearly identical between the two groups (50.79 vs. 50.73; P > 0.05).

Cross-classification by disease category and platform region likewise revealed no statistically significant regional differences within individual disease systems (oncology: P = 0.08; musculoskeletal diseases: P = 0.25; ophthalmology: P = 0.92).

Similarly, user engagement showed no significant association with information quality in the overall dataset (P = 0.669; Figure 4A) or after stratification by Chinese (P = 0.617; Figure 4B) and international platforms (P = 0.152; Figure 4C).

Collectively, these findings suggest that variation in video quality across disease categories may be smaller than differences observed across platform ecosystems. Nevertheless, because subgroup sample sizes were relatively limited, these findings should be interpreted with caution.

#### 3.2.3 Association Between Video Characteristics and DISCERN Scores

##### Video Duration

In univariable meta-regression, average video duration demonstrated a weak positive association with standardized DISCERN scores (β = 0.39; 95% CI: −0.04 to 0.82; P = 0.075).

However, after adjustment for platform region (Chinese vs. international), the association was no longer statistically significant. The regression coefficient was substantially attenuated after adjustment for platform region.

Exploratory disease-specific subgroup analyses demonstrated heterogeneous associations between video duration and standardized DISCERN scores. Negative associations were observed in oncology (β = −12.75, P < 0.001) and ophthalmology (β = −17.90, P = 0.014), whereas positive associations were found in cardiovascular diseases (β = 4.67, P = 0.033) and gastrointestinal diseases (β = 51.07, P < 0.001) (Figure 5).

However, leave-one-out sensitivity analyses showed that several associations lost statistical significance after exclusion of individual studies. For cardiovascular diseases and ophthalmology, statistical significance was no longer observed after sequential removal of individual studies. Although the associations remained significant for oncology and gastrointestinal diseases, these subgroup analyses were based on relatively small numbers of studies (n = 8 and n = 4, respectively), limiting the robustness of these findings.

##### User Engagement

No significant association was observed between the average number of likes and standardized DISCERN scores (P > 0.05).

Similar findings were observed across all subgroup analyses.

#### 3.2.4 Creator Professional Background

Meta-regression using the proportion of healthcare professionals as a continuous moderator demonstrated no significant association between creator professionalism and pooled DISCERN scores (P > 0.05) (Figure 6).

No statistically significant association was observed between the proportion of healthcare professionals and pooled DISCERN scores.

### 3.3 Multivariable Meta-regression Analysis

#### 3.3.1 Platform Region and Video Duration

To examine the fundamental characteristics of platform ecosystems, a multivariable meta-regression model incorporating platform region and average video duration was constructed. Average video duration was not significantly associated with standardized DISCERN scores (P = 0.891). Platform region showed a borderline association with standardized DISCERN scores (P = 0.058).

#### 3.3.2 Platform Region and Professional Participation

To further evaluate the independent effects of platform region and professional participation, a multivariable meta-regression model (Model A) was constructed using the proportion of healthcare professionals as a moderator. Collinearity diagnostics revealed a relatively strong negative correlation between platform region and professional density (r = −0.687).

After simultaneous adjustment for both variables, professional participation was not significantly associated with DISCERN scores (β = 0.091, 95% CI: −0.070 to 0.252, P = 0.268). In contrast, the effect of platform region remained significant after controlling for professional density (β = 9.962, 95% CI: 1.831 to 18.093, P = 0.016). After adjustment for professional participation, platform region remained significantly associated with standardized DISCERN scores, whereas professional participation did not.

As a comparison, Model B, which incorporated log-transformed average likes, showed that user popularity explained only a small proportion of the observed variance (R² = 3.29%), substantially less than the model including professional participation (R² = 9.06%).

#### 3.3.3 Residual Heterogeneity

Although the multivariable models identified platform region and professional participation as partial sources of heterogeneity, substantial residual heterogeneity remained. In Model A, residual heterogeneity was highly significant (Q_E = 6082.177, P < 0.001; residual I² = 99.08%). Overall, the model explained only 9.06% of the between-study variance, indicating that most variation in online health video quality remained unexplained by the variables included in the current analysis.

### 3.4 Assessment of Publication Bias and Sensitivity Analysis Qualitative and Quantitative Assessment of Bias

Visual inspection of the funnel plot for the primary outcome (standardized DISCERN score) demonstrated a largely symmetrical distribution. This observation was supported by Egger’s linear regression test, which did not detect significant publication bias or small-study effects (t = −1.26, P = 0.213).

Given the substantial clinical heterogeneity among included studies, additional stratified analyses were performed according to platform region to minimize potential masking effects caused by combining different digital ecosystems. Funnel plots for both Chinese domestic platforms and international platforms appeared approximately symmetrical around their respective pooled effect estimates. Egger’s tests likewise showed no significant asymmetry in either subgroup (Chinese platforms: t = 1.06, P = 0.299; international platforms: t = 0.99, P = 0.335).

To further evaluate the robustness of the pooled estimates, leave-one-out sensitivity analyses were conducted. Sequential exclusion of each individual study produced only minimal changes in the pooled standardized quality score, which remained within a narrow range of 47.32 to 50.62. No single study materially altered the pooled standardized DISCERN score.

Overall, neither publication bias nor substantial small-study effects were detected, and the leave-one-out analyses showed minimal variation in the pooled estimates.

## 4. Discussion

### 4.1 Principal Findings

This systematic review and meta-analysis synthesized empirical evidence from 88 cross-sectional studies evaluating a total of 18,688 videos, providing a comprehensive, global-scale assessment of online chronic disease health information across multiple digital platforms. Aggregated findings consistently demonstrate that the evidence-based quality of health communication across contemporary video-sharing ecosystems remains suboptimal. Validated appraisal instruments utilized in this study, such as the DISCERN criteria and JAMA benchmarks, evaluate not merely surface factual accuracy but also the systematic disclosure of clinical risks, alternative therapeutic options, and the transparency of evidence sources.

The pooled standardized scores across platforms hovered around the 50-point mark on a normalized 100-point scale. It is crucial to clarify that this threshold represents an investigator-defined benchmark calibrated from a clinical safety perspective rather than an absolute cutoff mandated by the official scales; nonetheless, this finding underscores that online medical videos systematically omit approximately half of the critical clinical context essential for safe patient self-management.

Although institutional warnings regarding digital health information governance on social media were articulated over a decade ago(13), video platforms remain an unreliable and highly volatile source of public health counseling. Even in the post-pandemic infodemic landscape, contemporary empirical evidence indicates that more than one-quarter of popular medical videos on mainstream platforms contain non-factual or misleading assertions(14).

Despite this suboptimal quality baseline, healthcare consumers continuously rely on online videos as a primary repository for medical information and actively utilize them as health decision-making tools (15). Within digital public health, "half-truth" narratives—which typically emphasize therapeutic efficacy while omitting adverse effects or contraindications—may compromise patient safety.

While this study sought to establish the first comprehensive global evidence map of chronic disease video quality, conventional content-level and creator-level variables—including specific disease categories, user engagement metrics (e.g., likes), and the proportion of medical professionals—yielded limited capacity to explain the observed quality variations. Consequently, the underlying determinants of online health information quality remain largely unexplained within traditional communication paradigms.

A robust finding from our subgroup analysis is that specific platforms, most notably YouTube, consistently demonstrated higher pooled quality scores compared to other ecosystems, suggesting that the current analytical framework may omit critical macro-level determinants of health video quality. As a major international platform operating outside domestic regulatory frameworks, YouTube’s superior performance might be attributed to its specialized institutional health information governance initiatives, such as formal source verification partnerships (16), though a definitive causal relationship cannot be established within the current cross-sectional design.

Our multivariable meta-regression analysis further indicated that geographic region maintained a robust, independent association with higher standardized DISCERN scores even after fully adjusting for physician involvement, whereas the density of certified medical professionals yielded no statistically significant regulatory effect on content quality. When contextualized alongside the non-significant impacts of disease categorization, video duration, and popularity metrics, these findings collectively demonstrate that micro-level creator or content attributes are insufficient to explain the variance in digital health information quality. Instead, unmeasured macro-level platform factors likely exert a more dominant influence.

Although geographic region was the only statistically significant moderator, the final multivariable model accounted for only 9.06% of the between-study variance. These findings suggest that the major determinants of online health information quality remain outside the variables examined in the current study.

Future infodemiological investigations must therefore transcend individualistic creator- or content-centric features and systematically examine structural platform-level mechanics, including algorithmic recommendation architectures, institutional information governance policies, and content moderation strategies.

### 4.2 Factors Underlying the Unpredictability of Video Quality

Traditional health communication frameworks assume that the quality of medical information is driven by supply-side factors and specific content characteristics. Specifically, conventional models assume that high-quality health education primarily originates from credentialed medical professionals, long-form narratives capable of providing comprehensive explanations, and the thorough evidence-based disclosures required by clinical domains with high patient vulnerability (17).

However, our multivariable meta-regression analysis substantially challenges these empirical assumptions. The proportion of physician creators, video duration, and clinical vulnerability parameters all failed to demonstrate robust, independent associations with standardized DISCERN scores, with the final model accounting for less than 10% of the total between-study heterogeneity. This pronounced explanatory deficit underscores a pivotal inquiry: why does the quality of online health information within short-video ecosystems remain so inherently unpredictable?

Our findings may be interpreted within a conceptual framework that we refer to as the Algorithmic Homogenization Hypothesis, a phenomenon partially corroborated in preliminary investigations(10). Short-form videos inherently simplify intricate medical topics into highly engaging multimedia formats by integrating visual, auditory, and textual modalities; yet, such compressed delivery systematically compromises information comprehensiveness and structural depth (18).

One plausible explanation is that the algorithmic recommendation mechanics governing short-video platforms exert a potent homogenizing force that systematically compresses the variance in quality metrics across distinct content typologies. Driven by an attention economy optimized for video completion rates, immediate emotional arousal, and rapid cognitive processing, platform architectures preferentially incentivize a standardized, algorithm-friendly baseline. Consequently, this structurally suppresses the nuanced, complex, and qualified language mandated by validated quality appraisal instruments.

This homogenization effect manifests across three distinct dimensions:

At the creator level, medical professionals often simplify narratives to align with platform engagement features, which can reduce the qualitative distinction between professional and non-professional content(19).

At the medium level, the attenuation of duration effects reflects how comprehensive educational content is structurally marginalized or fragmented into brief, micro-digestible segments, which systematically strips away the thorough risk disclosures and comparative treatment analyses required by clinical practice guidelines.

At the clinical level, the absence of pronounced disease-specific differentials suggests that macro platform-level communication attributes largely overshadow disease-specific infodemiological demands(20).

Ultimately, this pervasive algorithmic homogenization confines the vast majority of chronic disease health communication to a narrow performance band near the 50-point threshold on a normalized 0–100 scale. Within this mid-tier baseline, videos typically maintain superficial factual accuracy but systematically lack the advanced epistemological markers that characterize high-quality medical consensus, such as the disclosure of clinical uncertainties, balanced risk-benefit matrices, and rigorous evidence citations. This hypothesis may provide one possible explanation for both the relatively low pooled scores and the substantial residual heterogeneity.

When traditional predictors show limited explanatory power, the residual variance is more likely driven by unmeasured factors, such as localized platform policies or individual creator behaviors. Consequently, these findings suggest that variations in health video quality are more likely to reflect platform-specific environments than content or creator characteristics, prioritizing broad user engagement over clinical detail. This framework provides a clearer explanation for the varied findings currently reported in cross-sectional digital health studies.

### 4.3 Geographic Disparities in the Digital Health Infodemiological Landscape

A notable feature of this systematic review is the geographic concentration of the included literature. Of the 88 studies with identifiable institutional affiliations, 72 (82%) originated from mainland China, indicating that current evidence regarding online health video quality is primarily generated within a single national environment. This concentration has rarely been discussed in prior reviews of digital health communication.

Several factors may explain this concentration. Over the past decade, China experienced rapid growth in short-video adoption and digital health communication infrastructure (21).

Platforms like Douyin, Kwai, and Bilibili have accumulated large user bases and have become major sources for health information (22). This expansion has led to increased research attention regarding the reliability of digital medical information and its potential impact on public health. Additionally, this growth coincided with increased institutional support and academic interest in public health informatics within China, leading to a steady increase in published short-video studies.

Furthermore, this rapid expansion coincided with a substantial surge in institutional resource allocation and academic incentives within public health informatics and health communication domains in China, as evidenced by a steady bibliometric increase in short-video studies, where Douyin remains the most frequently appraised platform.

In contrast, despite the massive global active user bases of international platforms such as YouTube and TikTok, primary literature evaluating these ecosystems remains comparatively sparse within our synthesized database. This asymmetry should not be misinterpreted as a lack of scholarly interest outside of China, but rather as a reflection of divergent institutional research priorities. Existing Western literature heavily prioritizes traditional text-based digital portals, including patient portals, online health forums, and broader interactive digital health interventions, whereas systematic appraisals of short-form video health communication have only recently emerged. Contemporary reviews similarly indicate that global research on short-video health communication remains in its foundational stage and is characterized by pronounced methodological heterogeneity (23).

From a broader public health perspective, this geographic imbalance constitutes a critical empirical finding. Because platform governance frameworks, regulatory landscapes, recommendation algorithms, and native media ecologies vary significantly across jurisdictions, evidence derived predominantly from a single digital ecosystem may possess limited external generalizability. Therefore, future international collaborative efforts are urgently required to expand health video quality assessments into a wider array of national, linguistic, and cultural contexts, thereby establishing a globally representative evidence base for digital public health governance.

### 4.4 Methodological Implications: Appraisal Instruments in the Video and AI Eras

A critical methodological reflection arising from our analysis centers on whether traditional quality appraisal instruments, widely utilized within the health communication discipline, maintain robust construct validity when applied to contemporary multimedia platforms (24). For instance, the DISCERN instrument—the primary quantitative metric synthesized in this study—was developed by the British National Health Service (NHS) in 1999, an era structurally defined by printed patient information pamphlets.

In that historical communication landscape, health information dissemination relied almost exclusively on static, linear, and text-heavy media, where quality was epistemologically equated with textual completeness, such as the explicit listing of evidence sources, detailed explications of the risks of non-treatment, and a balanced presentation of alternative therapeutic regimens (25).

However, following nearly three decades of rapid technological evolution, the 2026 digital health communication matrix has thoroughly transitioned into a highly dynamic, multimodal tech-environment driven by artificial intelligence and video-centric architectures (26, 27). This paradigm shift from static paper-based text to the modern AI video era represents a fundamental restructuring of both content generation mechanisms and user-end cognitive perception loops. By reducing health communication quality to a rigid, one-dimensional disclosure checklist, traditional tools fail to encapsulate the distinctive operational parameters of modern short-video platforms.

In the developmental trajectory of digital health media, early video-sharing ecosystems, typified by YouTube, exhibited a primarily search-driven architecture. On these platforms, content traditionally leans toward medium- to long-form systematic expositions, providing creators with a sufficient temporal window to cite clinical guidelines, articulate complex pathophysiological mechanisms, and balance risk-benefit ratios. Consequently, traditional instruments preserve a relative degree of methodological validity when applied to search-driven architectures.

Conversely, the rapid rise of platforms like TikTok, Instagram Reels, Douyin, Kwai, and Rednote marks a transition to scroll-driven, immersive streaming architectures. The extreme temporal compression mandated by these platforms (frequently <60 seconds) and the severe algorithmic optimization for 3-second user retention rates compel medical professionals to decontextualize and downscale complex clinical knowledge. Within such algorithmic frameworks, it is physically and structurally unfeasible for creators to achieve the granular disclosures required by the DISCERN scale regarding long-term non-treatment outcomes or comprehensive alternative therapies.

The exhaustive criteria enforced by traditional text-based instruments are thus in structural conflict with the rapid, fragmented, and high-frequency attributes of contemporary short-video communication (28, 29). The actual public health efficacy of modern video content is heavily mediated by visual stratification, dynamic subtitling, creator charisma, and emotional resonance—multimodal variables that remain entirely unmeasured within 1999 instrument designs.

In conclusion, the applicability of traditional text-centric tools requires critical re-evaluation when confronting a 2026 infodemiological landscape populated by YouTube, TikTok, Reels, and artificial intelligence-generated content (AIGC). Continued reliance on obsolete instruments risks generating misleading public health governance conclusions by favoring formal checklist compliance over actual clinical reliability. The extreme residual heterogeneity (I^2^≈100%) observed in our meta-analysis partially reflects this fundamental limitation of current evaluation frameworks in capturing the critical features of modern digital health media.

These findings suggest that future public health informatics research would benefit from next-generation appraisal frameworks tailored to digital video environments. Future models should expand beyond static content scoring to integrate clinical accuracy, multimodal presentation characteristics, and platform-specific metrics.

### 4.5 Strengths of the Study

Compared with previous systematic reviews appraising online consumer health information, this study possesses several distinct methodological and conceptual advantages.

Comprehensive Cross-Ecosystem Evidence Mapping: To the authors’ knowledge, this is the first systematic review and meta-analysis specifically designed to synthesize the quality of online health videos concerning chronic disease management across both major international and Chinese digital platforms. Prior literature predominantly focused on isolated clinical subspecialties or single platforms, which constrained a holistic understanding of the macro digital health communication landscape. An increasing number of studies indicate that accessing health information online is a feasible way to assist patients with chronic diseases in long-term self-management. (30, 31). By aggregating empirical evidence from 88 primary studies involving 18,688 videos, this study delivers the most comprehensive infodemiological evidence map of chronic disease-related video content to date.

Cross-Ecosystem Comparison: A salient architectural advantage is the concurrent inclusion of both Chinese domestic (Bilibili, Douyin, Rednote, Kwai) and global international platforms (YouTube, TikTok), enabling a cross-ecosystem comparison rarely achieved in prior evidence syntheses. This cross-sectional design permits the identification of platform-level geographic variations that remain latent in single-platform investigations. Notably, although the review restricted its scope to literature utilizing English and Chinese database search strategies, congruent findings across other linguistic groups support the universal generalizability of the present conclusions(20, 32).

Clinical and Public Health Relevance: The deliberate focus on chronic conditions carries profound public health implications. Unlike acute or self-limiting conditions, which typically entail a brief information-seeking interval followed by definitive clinical resolution, chronic diseases necessitate sustained patient self-management over months or years. Consequently, healthcare consumers repetitively query online health repositories throughout their longitudinal disease trajectories, resulting in cumulative exposure to platform recommendation algorithms(33). Within this context, medical misinformation does not operate as an isolated, transient exposure; rather, its cognitive and behavioral effects accumulate over time, potentially compromising medication adherence, lifestyle modifications, healthcare utilization, and long-term clinical outcomes(34, 35). Therefore, evaluating video quality within chronic disease domains provides greater empirical relevance for public health policy than assessing short-term medical conditions.

Advanced Explanatory Modeling: Rather than merely reporting pooled quality estimates, this analysis utilizes multivariable meta-regression to systematically investigate the structural sources of between-study heterogeneity. By concurrently modeling platform region, video characteristics, creator professionalism, and audience engagement, the analysis demonstrates that conventional content-level indicators—including video duration, physician involvement, and popularity metrics—account for only a minimal proportion of the observed quality variation. This insight offers a critical direction for future digital health research to transcend traditional content-centric analyses and explore structural platform-level governance.

Methodological Standardization: The standardization of disparate quality appraisal instruments into a unified 0–100 scale successfully eliminates scaling bias across primary studies utilizing different measurement tools. This mathematical rescaling significantly elevates the interpretability and comparability of pooled estimates derived from heterogeneous evaluation criteria.

### 4.6 Limitations

Despite the rigorous methodological design, several structural limitations warrant prudent consideration.

Extreme Residual Heterogeneity: High residual heterogeneity was consistently observed across almost all consolidated analyses, with I² values approaching 100%. This high variance is structurally expected and clinically intelligible, given that online health videos differ fundamentally across platform architectures, clinical disciplines, source languages, creator attributes, recommendation algorithms, and appraisal tools. Consequently, although random-effects models and extensive subgroup stratifications were executed, the pooled estimates must be interpreted as aggregate average trends rather than universal characteristics representing all individual digital health assets.

Limited Predictability of Traditional Covariates: The multivariable meta-regression model accounted for only a modest fraction of the between-study heterogeneity (R² = 9.06%), demonstrating that approximately 91% of the quality variance is driven by unmeasured latent variables. These missing parameters likely involve platform-level macro factors—such as proprietary recommendation algorithms, backend moderation frameworks, and institutional content filtering policies—that are inherently uncaptured in primary cross-sectional literatures. While this unexplained variation restricts the mathematical precision of the pooled estimates, it accurately reflects the real-world diversity and chaotic dynamics of the contemporary digital health ecosystem.

Statistical Power of Bias Diagnostics: Although the funnel plots and Egger’s linear regression tests yielded no statistically significant evidence of publication bias, these diagnostics require cautious interpretation. In single-arm meta-analyses evaluating continuous outcomes, funnel plot asymmetry frequently reflects genuine clinical or methodological heterogeneity among studies rather than selective non-publication of null results. Conversely, the presence of extreme residual heterogeneity can actively attenuate the statistical power of conventional regression tests to detect subtle small-study effects. Therefore, the absence of statistical significance should not be construed as definitive proof of the complete absence of publication bias.

Anachronistic Limitations of Appraisal Tools: A fundamental limitation relates to the intrinsic constraints of the appraisal instruments themselves. The majority of the included primary studies relied on the DISCERN, GQS, or JAMA benchmarks, which were originally conceptualized to evaluate written consumer health materials or printed pamphlets during the late 1990s. Although these instruments remain the most widely accepted frameworks within contemporary evidence synthesis, they are structurally ill-equipped to capture the defining characteristics of modern social media platforms, including algorithmic amplification, visual persuasion mechanics, real-time user-producer interactions, and push-based recommendation dynamics.

Database and Subgroup Sample Constraints: This review exclusively included peer-reviewed literature indexed in major English-language databases, which may inherently omit relevant gray literature, institutional white papers, or studies published in other localized national languages. Additionally, certain disease-specific subgroup analyses contained a relatively sparse number of primary studies, which restricted statistical power. Given that leave-one-out sensitivity analyses indicated that several seemingly significant subgroup associations were heavily driven by individual influential studies, these exploratory findings must be interpreted with caution and warrant further validation through large-scale primary investigations.

## 5. Conclusions

This systematic review and meta-analysis provides the most comprehensive evidence synthesis to date regarding the quality of online health videos related to chronic conditions across major Chinese domestic and international platforms. Despite the rapid expansion of digital health communication, the overall evidence-based quality of publicly available video assets remains structurally suboptimal when measured against established appraisal instruments.

Crucially, the empirical findings demonstrate that traditional content- and creator-level predictors—including creator professional credentials, video duration, specific disease categories, and audience engagement metrics—possess limited capacity to explain the variance in video quality. Although platform region emerged as the sole statistically significant moderator, the final multivariable meta-regression model accounted for only a minor fraction of the observed between-study heterogeneity, indicating that the predominant drivers of health information quality remain largely unexplained within current evaluation frameworks.

These insights highlight a critical pivot for future infodemiological and public health informatics research. Rather than focusing exclusively on individual creator behaviors or micro-level content features, future investigations must systematically examine macro platform-level mechanisms. Specifically, exploring proprietary recommendation algorithms, institutional content governance strategies, and algorithmic moderation policies is essential, as these systemic variables likely exert a more decisive influence on the baseline quality of health information presented to end-users.

Furthermore, in a contemporary ecosystem dominated by algorithm-driven short videos and artificial intelligence-generated content (AIGC), the widespread and continued reliance on anachronistic, text-centric appraisal tools, such as the DISCERN criteria, GQS, and JAMA benchmarks, warrants critical reconsideration. The development of next-generation evaluation frameworks that dynamically integrate clinical accuracy, multimodal audiovisual communication traits, and platform ecologies is vital to accurately appraise digital health information and support evidence-based public health governance.

Ultimately, the empirical evidence underscores the necessity of a fundamental paradigm shift from a creator-centric explanatory model to a platform-centric architectural framework to robustly understand and regulate the quality landscape of online health information.

## Supporting information

PRISMA_2020_checklist

Supplementary Appendix 1 Full Search Strategies

Supplementary Appendix 1 Full Search Strategies

Table 2 Overall video quality analysis results

Table 3 Subgroup Analysis of DISCERN by Platform Region

Table 4 Subgroup Analysis of DISCERN by Disease System

## Data Availability Statement

The datasets generated and analyzed during the current study are available from the corresponding author on reasonable request.

## Abbreviations

AIGC: Artificial Intelligence-Generated Content
CDC: Centers for Disease Control and Prevention
CHE: Catastrophic Health Expenditure
CI: Confidence Interval
DALYs: Disability-Adjusted Life Years
GQS: Global Quality Scale
IQR: Interquartile Range
JAMA: Journal of the American Medical Association
JBI: Joanna Briggs Institute
MRAW: Raw Mean
NHS: National Health Service
PI: Prediction Interval
PICO: Population, Intervention, Comparison, and Outcome
PRISMA: Preferred Reporting Items for Systematic Reviews and Meta-Analyses
PROSPERO: International Prospective Register of Systematic Reviews
REML: Restricted Maximum Likelihood
SD: Standard Deviation
WHO: World Health Organization

---

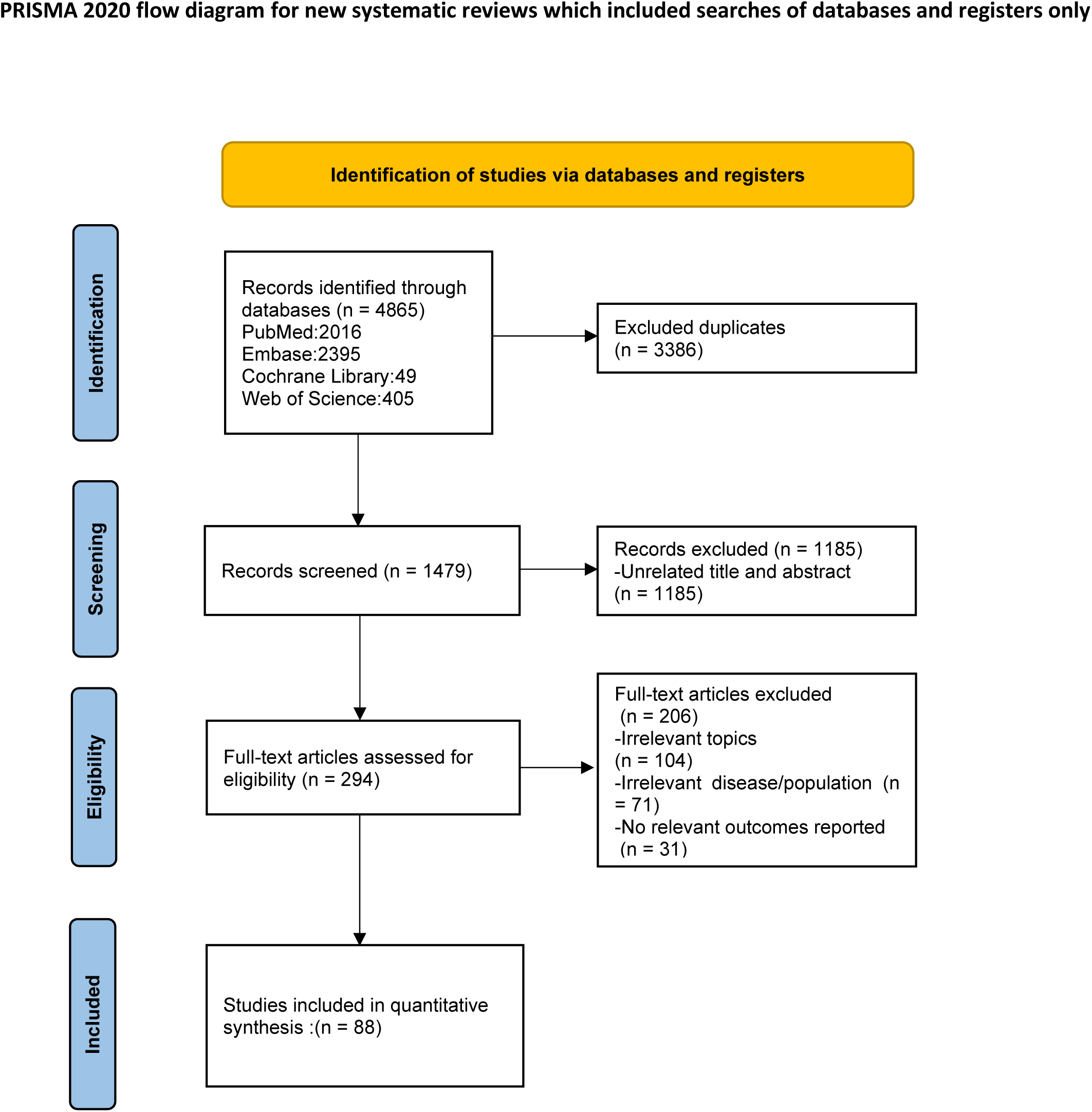

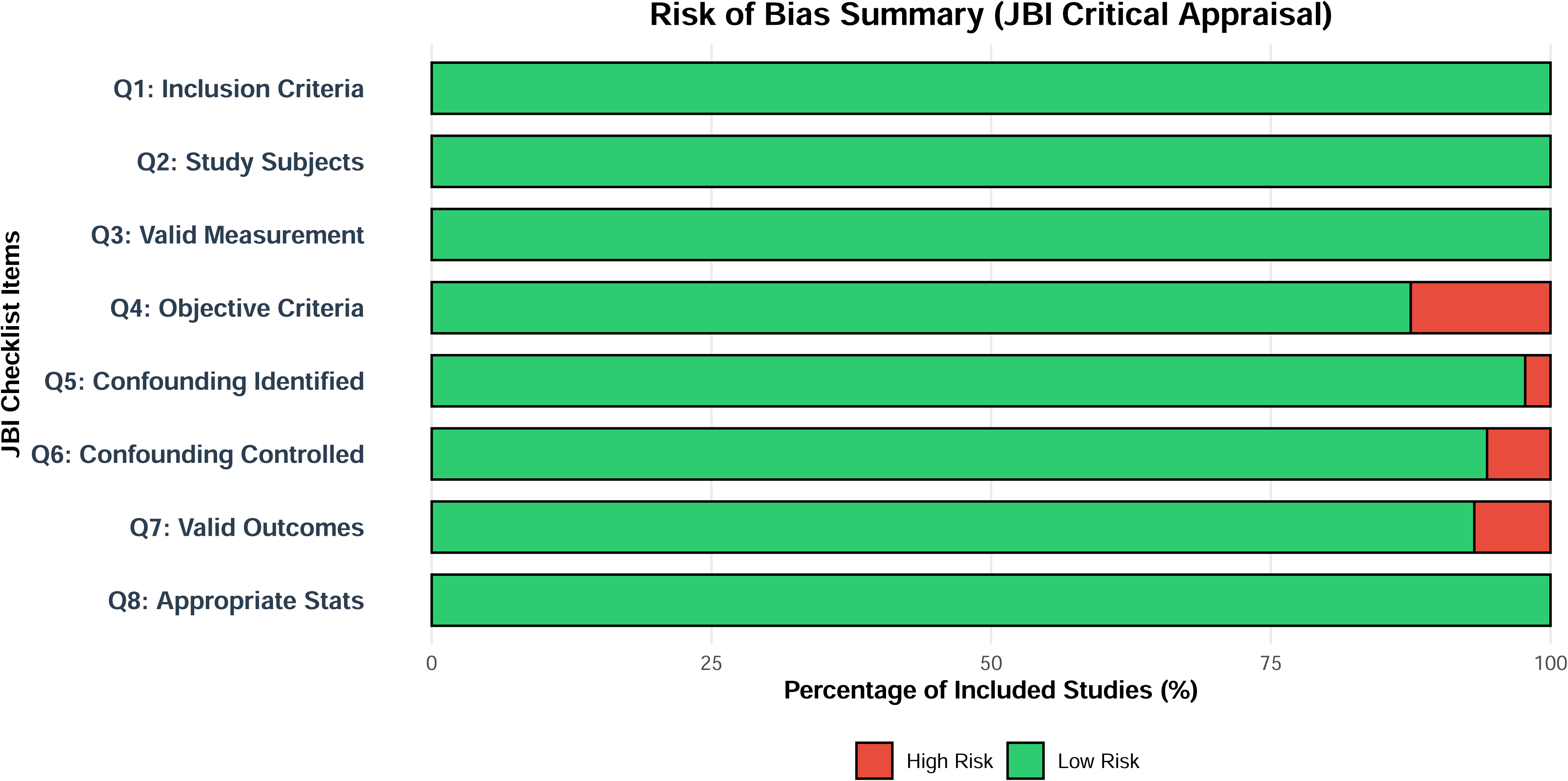

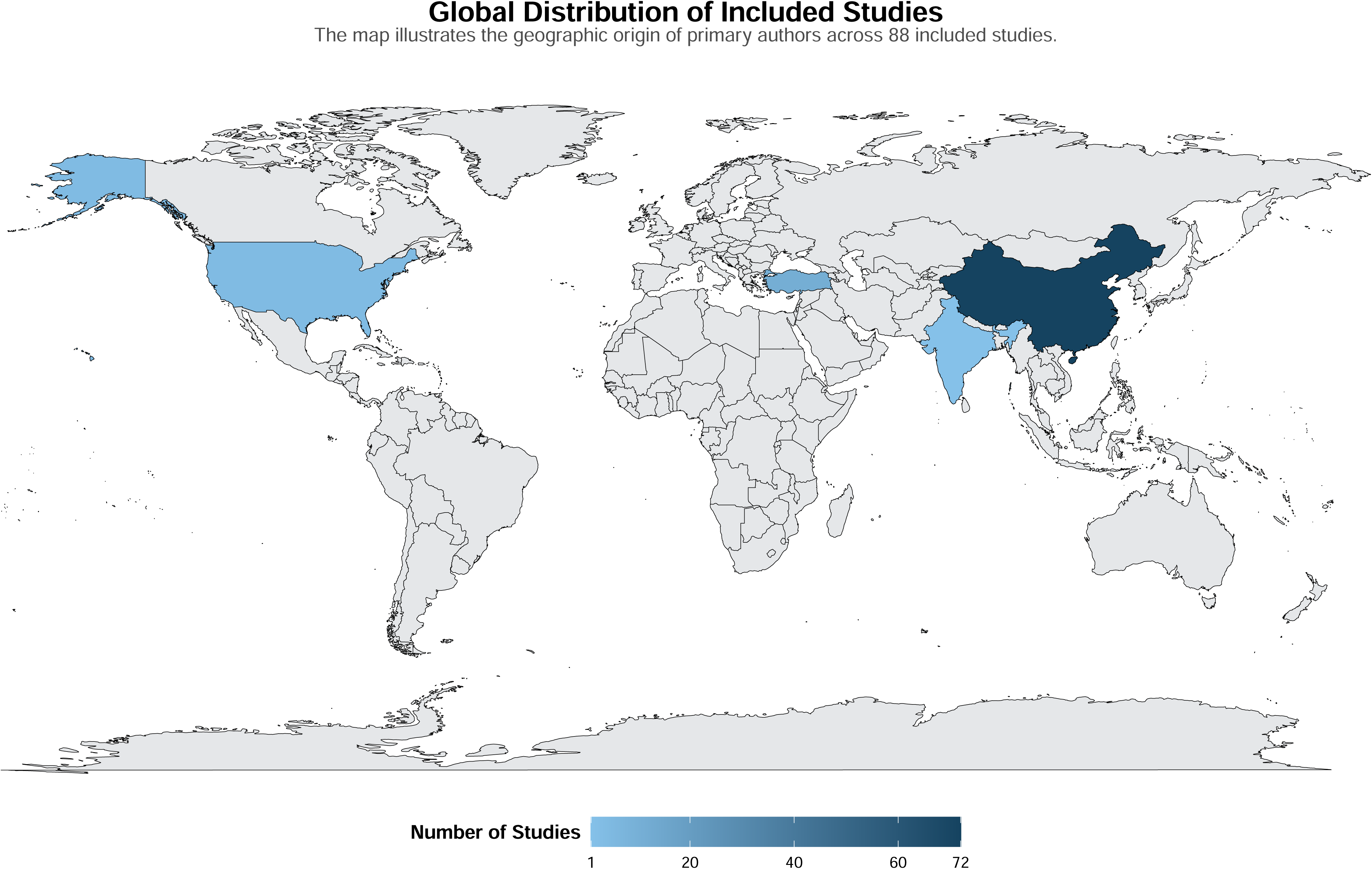

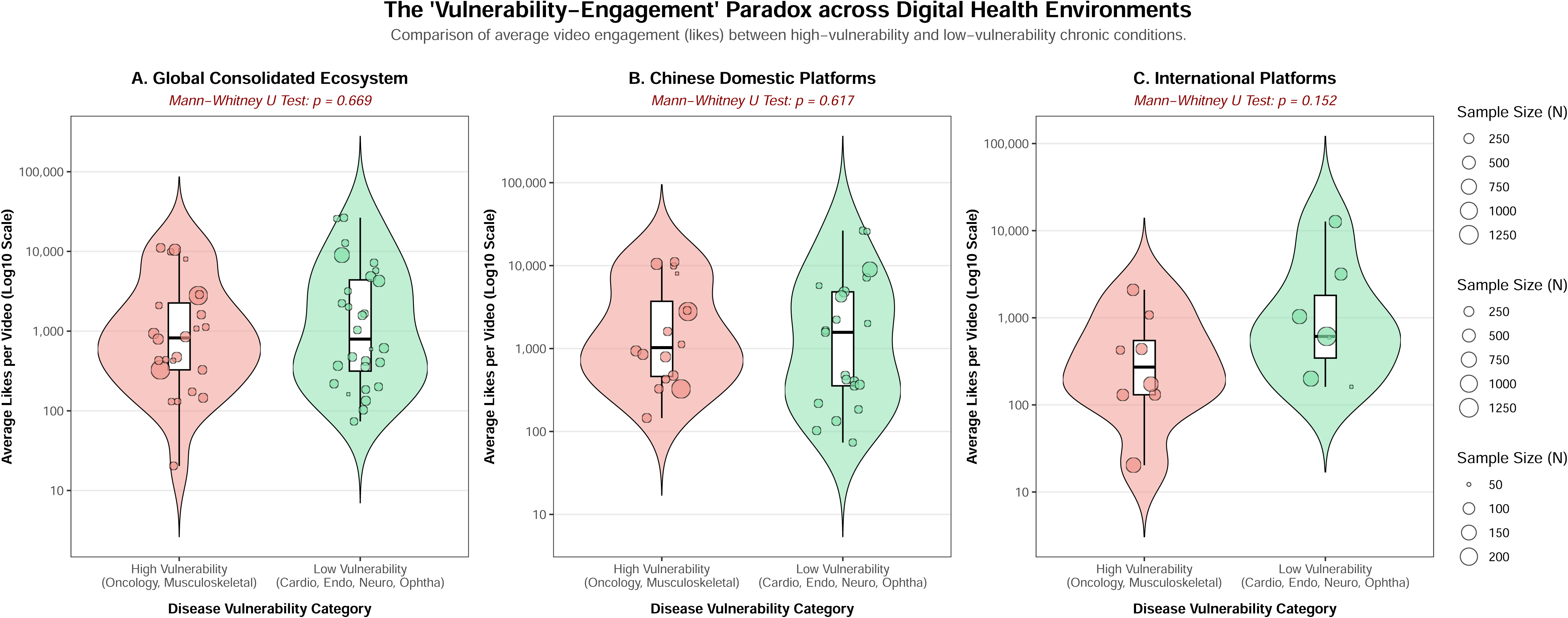

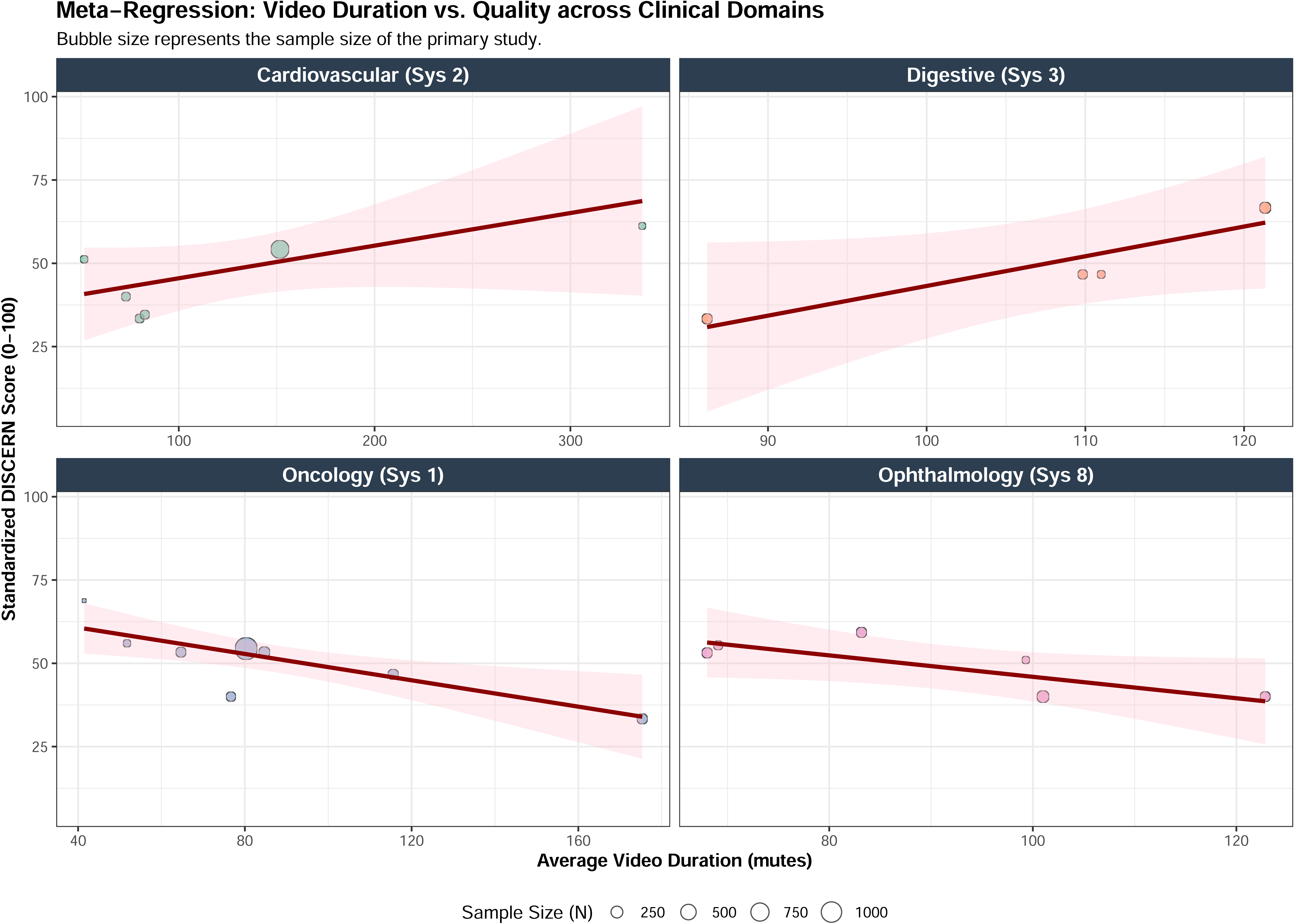

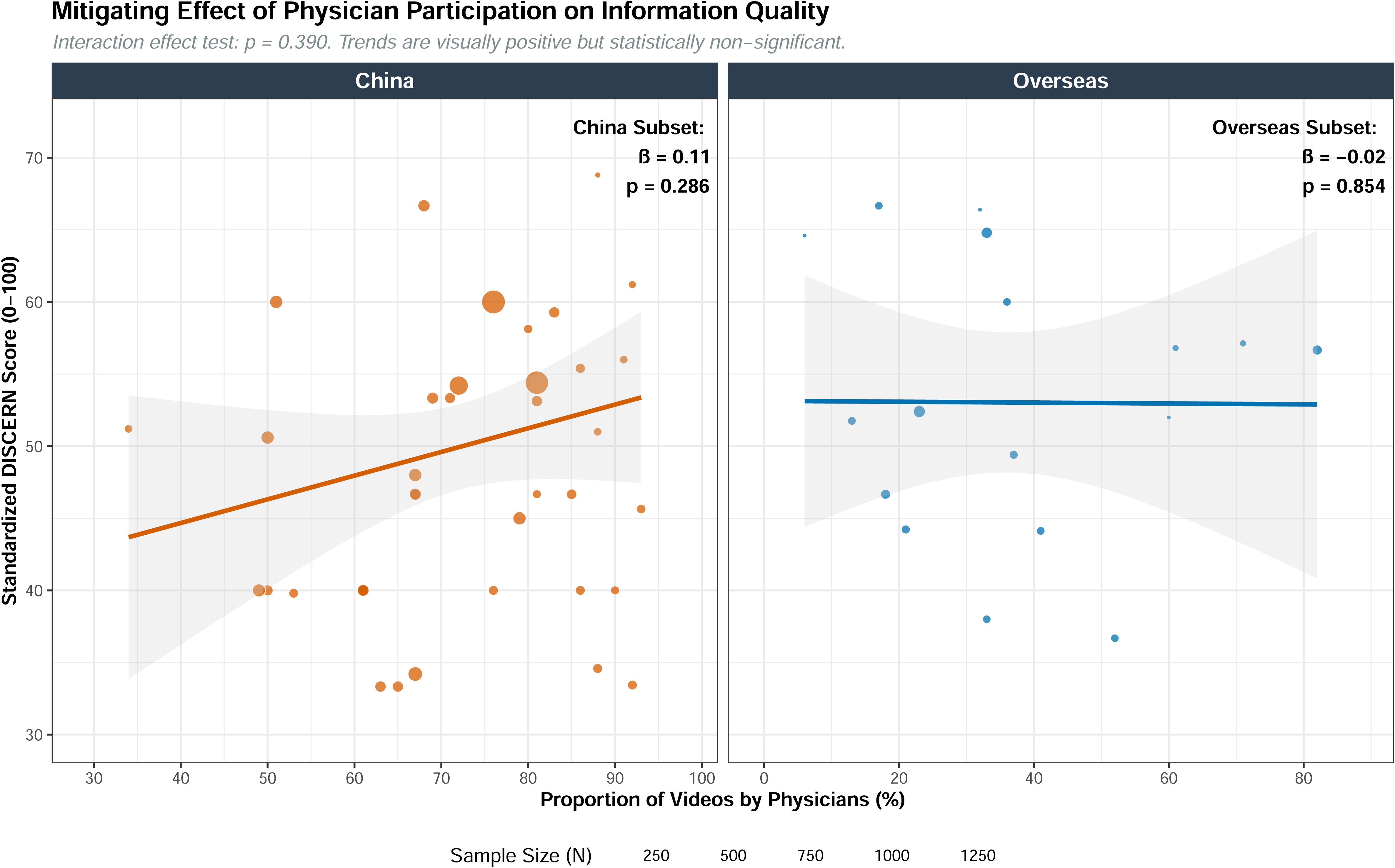

