## Supplementary Appendix 1 Full Search Strategies for "Quality of Chronic Disease–Related Health Videos Across Social Media Platforms: A Systematic Review and Meta-analysis"

The complete electronic search strategies used for all databases are provided below. Searches were conducted from database inception to April 30, 2026.

**Pubmed:**

(

"Bilibili"[Title/Abstract]

OR "TikTok"[Title/Abstract]

OR "Douyin"[Title/Abstract]

OR "Kuaishou"[Title/Abstract]

OR "Kwai"[Title/Abstract]

OR "short video*"[Title/Abstract]

OR "video-sharing platform*"[Title/Abstract]

OR "Social Media"[MeSH] OR "social media"[Title/Abstract]

OR "social media video*"[Title/Abstract]

)

AND

(

"health information"[Title/Abstract]

OR "medical information"[Title/Abstract]

OR "health education"[Title/Abstract]

OR "patient education"[Title/Abstract]

OR "health communication"[Title/Abstract]

OR "medical content*"[Title/Abstract]

OR "Consumer Health Information"[MeSH] OR "consumer health information"[Title/Abstract]

OR "Infodemiology"[Title/Abstract]

)

AND

(

"quality"[Title/Abstract]

OR "reliability"[Title/Abstract]

OR "accuracy"[Title/Abstract]

OR "completeness"[Title/Abstract]

OR "DISCERN"[Title/Abstract]

OR "Global Quality Scale"[Title/Abstract]

OR "GQS"[Title/Abstract]

OR "JAMA benchmark*"[Title/Abstract]

OR "Quality of Information"[MeSH] OR "information quality"[Title/Abstract]

)

**Embase:**
(

'bilibili':ti,ab OR 'tiktok':ti,ab OR 'douyin':ti,ab OR 'kuaishou':ti,ab OR 'kwai':ti,ab OR 'short video*':ti,ab OR 'video sharing platform*':ti,ab OR 'social media'/exp OR 'social media video*':ti,ab

)

AND

(

'health information':ti,ab OR 'medical information':ti,ab OR 'health education':ti,ab OR 'patient education':ti,ab OR 'health communication':ti,ab OR 'medical content*':ti,ab OR 'consumer health information'/exp OR 'infodemiology':ti,ab

)

AND

(

'quality':ti,ab OR 'reliability':ti,ab OR 'accuracy':ti,ab OR 'completeness':ti,ab OR 'discern':ti,ab OR 'global quality scale':ti,ab OR 'gqs':ti,ab OR 'jama benchmark*':ti,ab OR 'information quality'/exp

)

**Cochrane Library:**
(

[mh "Social Media"]

OR Bilibili:ti,ab,kw

OR TikTok:ti,ab,kw

OR Douyin:ti,ab,kw

OR Kuaishou:ti,ab,kw

OR Kwai:ti,ab,kw

OR short NEXT video*:ti,ab,kw

OR video NEXT platform*:ti,ab,kw

)

AND

(

[mh "Consumer Health Information"]

OR "health information":ti,ab,kw

OR "medical information":ti,ab,kw

OR "health education":ti,ab,kw

OR "patient education":ti,ab,kw

OR medical NEXT content*:ti,ab,kw

OR Infodemiology:ti,ab,kw

)

AND

(

[mh "Quality of Information"]

OR quality:ti,ab,kw

OR reliability:ti,ab,kw

OR accuracy:ti,ab,kw

OR completeness:ti,ab,kw

OR DISCERN:ti,ab,kw

OR GQS:ti,ab,kw

OR JAMA NEXT benchmark*:ti,ab,kw

)

**WOS:**
TS=(

("Bilibili" OR "TikTok" OR "Douyin" OR "Kuaishou" OR "Kwai" OR "short video*" OR "video-sharing platform*" OR "social media video*")

AND

("health information" OR "medical information" OR "health education" OR "patient education" OR "health communication" OR "medical content*" OR "Infodemiology" OR "Consumer Health Information")

AND

("quality" OR "reliability" OR "accuracy" OR "completeness" OR "transparency" OR "DISCERN" OR "Global Quality Scale" OR "GQS" OR "JAMA benchmark*")

)
