## Supplementary Appendix 1 Full Search Strategies for "Quality of Chronic Disease–Related Health Videos Across Social Media Platforms: A Systematic Review and Meta-analysis"

**Table 1. Characteristics of included studies.**

| Study (Year) & Country | | | Platform Ecosystem | Clinical Domain (Specific Disease) | Search Period | Videos(N) | | Creator Profile (Physician Proportion, %) | Engagement Metrics (Mean Likes) | | Quality Assessment Tools | Risk of Bias |
| --- | --- | --- | --- | --- | --- | --- | --- | --- | --- | --- | --- | --- |
|  | W. Zhu et al. (2025), China | Bilibili, Douyin, Kwai | | Oncology (Esophageal cancer) | NR | 311 | NR | | | NR | NR | Low |
|  | X. Zheng et al. (2025), China | Douyin, Rednote, Kwai | | Oncology (Thyroid cancer) | 2023-2026 | 1248 | 81 | | | 2793.26 | DISCERN, GQS, JAMA | Low |
|  | J. Peng et al. (2025), China | Bilibili, Douyin, Kwai | | Oncology (Pancreatic cancer) | 2023-2026 | 300 | 79 | | | 936.00 | DISCERN, GQS | Low |
|  | J. Zhang et al. (2024), China | Bilibili, Kwai | | Oncology (Cervical cancer) | 2023-2026 | 163 | 61 | | | 2876.67 | DISCERN, GQS | Low |
|  | Q. Ding et al. (2025), China | Douyin | | Oncology (Esophageal cancer) | 2023-2026 | 246 | 86 | | | NR | NR | Low |
|  | S. Yang et al. (2023), China | Douyin | | Oncology (Thyroid cancer) | 2023-2026 | 100 | 91 | | | 9899.76 | DISCERN, GQS | Low |
|  | H. Liu et al. (2024), China | Bilibili, Douyin | | Oncology (Breast cancer) | 2020-2022 | 200 | NR | | | 11092.33 | DISCERN, GQS | Low |
|  | L. Wang et al. (2023), China | Douyin | | Oncology (Thyroid cancer) | 2023-2026 | 56 | 88 | | | 8020.61 | DISCERN | Low |
|  | R. Zhang et al. (2024), China | Bilibili, Douyin | | Oncology (Brain tumor) | 2020-2022 | 200 | NR | | | NR | NR | Low |
|  | X. Zhao et al. (2024), China | Douyin, Kwai | | Oncology (Lung cancer) | 2023-2026 | 186 | 71 | | | 1605.33 | DISCERN, GQS | Low |
|  | S. Zheng et al. (2023), China | Bilibili, Douyin | | Oncology (Liver cancer) | 2023-2026 | 200 | NR | | | NR | NR | Low |
|  | X. Zheng et al. (2025), China | Douyin, Rednote, Kwai | | Oncology (Lung cancer) | 2023-2026 | 1288 | 76 | | | 326.08 | GQS, JAMA | Low |
|  | M. Liang et al. (2025), China | Bilibili, Douyin | | Oncology (Prostate cancer) | 2023-2026 | 182 | NR | | | NR | NR | Low |
|  | L. Ren et al. (2025), China | Douyin | | Oncology (Melanoma) | 2023-2026 | 113 | 90 | | | 1123.33 | NR | Low |
|  | J. Su et al. (2026), China | Bilibili, Douyin | | Oncology (Renal cell carcinoma) | 2023-2026 | 196 | 65 | | | 326.83 | DISCERN, GQS | Low |
|  | L. Su et al. (2025), China | Bilibili, Douyin, Kwai | | Oncology (Cervical cancer) | 2023-2026 | 259 | NR | | | 472.50 | NR | Low |
|  | H. Sha et al. (2026), China | Bilibili, Douyin | | Oncology (Hodgkin lymphoma) | 2023-2026 | 225 | 69 | | | 145.33 | DISCERN, GQS, JAMA | Low |
|  | X. Zhou et al. (2026), China | Douyin, Rednote | | Oncology (Ovarian cancer) | 2023-2026 | 155 | NR | | | NR | NR | Low |
|  | Z. Wang et al. (2026), China | Bilibili, Douyin, Kwai | | Oncology (Lung cancer) | 2023-2026 | 300 | 42 | | | NR | NR | Low |
|  | Z. Liu et al. (2024), China | Youtube | | Oncology (Laryngeal carcinoma) | 2023-2026 | 99 | 17 | | | 131.00 | DISCERN, GQS | Low |
|  | Q. Zhang et al. (2025), China | Youtube | | Oncology (Oral cancer) | 2023-2026 | 150 | 82 | | | 173.10 | DISCERN | Low |
|  | Y. Liu et al. (2026), China | Youtube | | Oncology (Cervical cancer) | 2023-2026 | 70 | 61 | | | 425.42 | DISCERN, GQS, JAMA | Low |
|  | Z. Chen et al. (2023), China | Youtube | | Oncology (Anal cancer) | 2023-2026 | 105 | 13 | | | 130.4 | DISCERN, GQS, JAMA | Low |
|  | J. Wu et al. (2025), China | Douyin | | Cardiovascular (Hypertension) | 2020-2022 | 139 | 86 | | | 26433.00 | GQS | Low |
|  | Z. Wang et al. (2026), China | Douyin | | Cardiovascular (Stroke) | 2023-2026 | 100 | 34 | | | 25798.33 | DISCERN, GQS | Low |
|  | P. Hao et al. (2025), China | Douyin | | Cardiovascular (Coronary artery disease) | 2023-2026 | 122 | 80 | | | NR | DISCERN | Low |
|  | N. Cui et al. (2023), China | Douyin | | Cardiovascular (Mitral valve regurgitation) | 2023-2026 | 88 | 92 | | | 5728.02 | DISCERN, GQS, JAMA | Low |
|  | X. Gong et al. (2023), China | Douyin | | Cardiovascular (Heart failure) | 2023-2026 | 141 | 92 | | | 1655.67 | DISCERN | Low |
|  | R. Ge et al. (2025), China | Bilibili, Douyin | | Cardiovascular (Stroke) | 2023-2026 | 142 | NR | | | NR | NR | Low |
|  | X. Gong et al. (2024), China | Douyin | | Cardiovascular (Coronary heart disease) | 2023-2026 | 145 | 88 | | | 7198.33 | DISCERN, GQS | Low |
|  | Q. Li et al. (2025), China | Bilibili, Douyin, Rednote, Kwai | | Cardiovascular (Atherosclerosis) | 2020-2022 | 764 | 72 | | | 8993.23 | DISCERN, GQS, JAMA | Low |
|  | J. Wang et al. (2025), China | Bilibili, Douyin | | Cardiovascular (Abdominal aortic aneurysm) | 2023-2026 | 140 | 76 | | | 73.58 | DISCERN, GQS | Low |
|  | H. Shu et al. (2026), China | Youtube | | Cardiovascular (Stroke) | 2023-2026 | 100 | 36 | | | NR | DISCERN, GQS, JAMA | Low |
|  | E. Yılmaz et al. (2024), Turkey | Youtube | | Cardiovascular (Heart failure) | 2023-2026 | 162 | NR | | | 200.00 | DISCERN, GQS | Low |
|  | G. Jammula et al. (2025), India | Youtube | | Cardiovascular (Hypertension) | 2023-2026 | 95 | 33 | | | NR | NR | Low |
|  | X. Yin et al. (2025), China | Bilibili, Douyin | | Digestive (Chronic pancreatitis) | 2023-2026 | 112 | 81 | | | 166.83 | DISCERN, GQS | Low |
|  | Y. Liang et al. (2024), China | Bilibili, Douyin | | Digestive (Gastroesophageal reflux disease) | 2023-2026 | 156 | NR | | | NR | NR | Low |
|  | Y. Cai et al. (2024), China | Bilibili, Douyin | | Digestive (Gastroesophageal reflux disease) | 2023-2026 | 164 | 85 | | | 1120.50 | DISCERN, GQS | Low |
|  | T. Chen et al. (2024), China | Bilibili, Douyin, Kwai | | Digestive (Irritable bowel syndrome) | 2023-2026 | 244 | 68 | | | 220.58 | DISCERN, GQS, JAMA | Low |
|  | J. Cai et al. (2026), China | Douyin, Rednote | | Digestive (Crohn's disease) | 2023-2026 | 200 | 63 | | | 282.67 | DISCERN, JAMA | Low |
|  | G. Waidyaratne et al. (2024), USA | Tiktok | | Digestive (Irritable bowel syndrome) | 2023-2026 | 100 | 33 | | | NR | DISCERN | Low |
|  | S. Lin et al. (2025), China | Bilibili, Douyin | | Endocrine and Metabolic (Diabetic kidney disease) | 2023-2026 | 200 | NR | | | NR | NR | Low |
|  | Y. Chen et al. (2024), China | Bilibili, Douyin | | Endocrine and Metabolic (Thyroid nodules) | 2023-2026 | 200 | NR | | | NR | NR | Low |
|  | Q. Cai et al. (2024), China | Bilibili, Douyin | | Endocrine and Metabolic (Gestational diabetes) | 2023-2026 | 135 | 53 | | | 2230.15 | DISCERN, GQS, JAMA | Low |
|  | R. Ding et al. (2024), China | Bilibili, Douyin, Kwai | | Endocrine and Metabolic (Steatotic liver disease) | 2020-2022 | 198 | NR | | | NR | NR | Low |
|  | K. Sun et al. (2025), China | Bilibili, Douyin | | Endocrine and Metabolic (Diabetes) | 2023-2026 | 300 | 51 | | | NR | DISCERN, JAMA | Low |
|  | R. Xu et al. (2025), China | Bilibili, Douyin | | Endocrine and Metabolic (Premature ovarian failure) | 2023-2026 | 187 | 61 | | | 102.67 | DISCERN, GQS | Low |
|  | H. Yan et al. (2026), China | Bilibili, Douyin, Rednote | | Endocrine and Metabolic (Hyperlipidemia) | 2023-2026 | 233 | 67 | | | 367.00 | DISCERN, GQS, JAMA | Low |
|  | A. Shah et al. (2025), USA | Tiktok | | Endocrine and Metabolic (Hyperthyroidism) | 2023-2026 | 115 | 37 | | | 12725 | DISCERN, GQS | Low |
|  | S. Wang et al. (2025), China | Youtube | | Endocrine and Metabolic (Hashimoto's thyroiditis) | 2023-2026 | 200 | NR | | | NR | DISCERN, JAMA | Low |
|  | H. Zeng et al. (2025), China | Bilibili, Douyin | | Renal and Urinary (Uraemia) | 2023-2026 | 153 | NR | | | 5276.67 | GQS, | Low |
|  | B. Aktas et al. (2023), Turkey | Youtube | | Renal and Urinary (Prostatitis) | 2023-2026 | 200 | 33 | | | 224.63 | DISCERN, GQS, JAMA | Low |
|  | H. Zhang et al. (2025), China | Douyin | | Musculoskeletal (Osteoporosis) | 2020-2022 | 128 | 93 | | | NR | DISCERN | Low |
|  | Q. Zuo et al. (2025), China | Bilibili, Douyin | | Musculoskeletal (Osteoarthritis) | 2023-2026 | 189 | NR | | | NR | NR | Low |
|  | Y. Zhang et al. (2025), China | Douyin, Rednote | | Musculoskeletal (Osteoarthritis) | 2023-2026 | 300 | NR | | | NR | NR | Low |
|  | X. Duan et al. (2026), China | Bilibili, Douyin, Rednote | | Musculoskeletal (Knee osteoarthritis) | 2023-2026 | 300 | 50 | | | 850.00 | DISCERN, GQS, JAMA | Low |
|  | J. Tu et al. (2025), China | Bilibili, Douyin | | Musculoskeletal (Knee osteoarthritis) | 2023-2026 | 164 | 80 | | | 428.33 | NR | Low |
|  | Y. Wang et al. (2025), China | Bilibili, Douyin, Rednote, Kwai | | Musculoskeletal (Lumbar disc herniation) | 2023-2026 | 384 | 67 | | | 10477.67 | DISCERN, GQS | Low |
|  | B. Wei et al. (2026), China | Bilibili, Douyin | | Musculoskeletal (Osteonecrosis) | 2023-2026 | 162 | NR | | | NR | NR | Low |
|  | R. Liu et al. (2026), China | Bilibili, Douyin | | Musculoskeletal (Ankylosing spondylitis) | 2023-2026 | 200 | 87 | | | NR | NR | Low |
|  | X. Wei et al. (2026), China | Bilibili, Douyin, Rednote | | Musculoskeletal (Ankylosing spondylitis) | 2023-2026 | 300 | 67 | | | 793.67 | DISCERN, GQS, JAMA | Low |
|  | T. Q. Tabarestani et al. (2023), USA | Tiktok | | Musculoskeletal (Achilles tendinopathy) | 2023-2026 | 100 | 52 | | | 438.20 | DISCERN | Low |
|  | M. E. Onder et al. (2021), Turkey | Youtube | | Musculoskeletal (Psoriatic arthritis) | 2020-2022 | 155 | NR | | | 20.33 | DISCERN, GQS, JAMA | Low |
|  | J. M. Azarabadi et al. (2025), Turkey | Youtube | | Musculoskeletal (Osteonecrosis) | 2020-2022 | 70 | 71 | | | 1076 | DISCERN, GQS, JAMA | Low |
|  | M. E. Onder et al. (2022), Turkey | Youtube | | Musculoskeletal (Osteoporosis) | 2023-2026 | 238 | NR | | | NR | DISCERN, GQS | Low |
|  | M. G. Simsek et al. (2026), Turkey | Youtube | | Musculoskeletal (Cervical disc herniation) | 2020-2022 | 104 | 41 | | | 2096.4 | DISCERN, GQS, JAMA | Low |
|  | B. Jawich et al. (2026), USA | Tiktok | | Musculoskeletal (Shoulder injuries) | 2023-2026 | 209 | 20 | | | NR | NR | Low |
|  | X. Qi et al. (2025), China | Bilibili, Douyin, Rednote, Kwai | | Neurological and Psychiatric (Bipolar disorder) | 2023-2026 | 606 | NR | | | NR | NR | Low |
|  | G. Wang et al. (2025), China | Bilibili, Douyin | | Neurological and Psychiatric (Adolescent depression) | 2023-2026 | 188 | NR | | | NR | NR | Low |
|  | S. Qian et al. (2026), China | Bilibili, Douyin | | Neurological and Psychiatric (Migraine) | 2023-2026 | 175 | 67 | | | 1570.17 | DISCERN, GQS | Low |
|  | Q. Wang et al. (2026), China | Bilibili, Douyin, Rednote, Kwai | | Neurological and Psychiatric (Sleep disorders) | 2023-2026 | 400 | 64 | | | 4259.33 | NR | Low |
|  | Y. Zhang et al. (2025), China | Bilibili, Douyin, Kwai | | Neurological and Psychiatric (Spinal cord injury) | 2023-2026 | 251 | NR | | | NR | NR | Low |
|  | Q. Liao et al. (2026), China | Bilibili, Douyin | | Neurological and Psychiatric (Myasthenia gravis) | 2023-2026 | 225 | 61 | | | 133.67 | GQS | Low |
|  | M. Alpua et al. (2026), Turkey | Youtube | | Neurological and Psychiatric (Functional neurological disorder) | 2023-2026 | 50 | 6 | | | 594.56 | DISCERN, GQS, JAMA | Low |
|  | A. E. Kaya et al. (2023), Turkey | Youtube | | Neurological and Psychiatric (Agoraphobia) | 2023-2026 | 50 | 32 | | | NR | DISCERN, GQS | Low |
|  | X. Liu et al. (2026), China | Youtube | | Neurological and Psychiatric (Parkinson's disease) | 2023-2026 | 147 | 18 | | | 1033.33 | DISCERN, GQS | Low |
|  | H. Wang et al. (2025), China | Bilibili, Douyin | | Ophthalmology (Thyroid eye disease) | 2023-2026 | 152 | 86 | | | 184.83 | DISCERN, GQS | Low |
|  | M. Liu et al. (2026), China | Bilibili, Douyin | | Ophthalmology (Amblyopia) | 2023-2026 | 185 | 50 | | | 218.17 | DISCERN, GQS, JAMA | Low |
|  | M. Huang et al. (2025), China | Douyin | | Ophthalmology (Dry eye) | 2023-2026 | 199 | 81 | | | 405.00 | DISCERN | Low |
|  | L. Jiang et al. (2026), China | Douyin | | Ophthalmology (Age-related macular) | 2023-2026 | 196 | 83 | | | 474.08 | DISCERN | Low |
|  | L. Jiang et al. (2025), China | Douyin | | Ophthalmology (Diabetic retinopathy) | 2023-2026 | 200 | 87 | | | 422.58 | NR | Low |
|  | M. Li et al. (2025), China | Bilibili, Douyin, Kwai | | Ophthalmology (Myopia) | 2023-2026 | 284 | 49 | | | 4822.50 | DISCERN, GQS | Low |
|  | J. Cao et al. (2025), China | Douyin | | Ophthalmology (Cataract) | 2023-2026 | 100 | 88 | | | 2009.1 | DISCERN, GQS, JAMA | Low |
|  | Y. Wang et al. (2024), China | Douyin | | Ophthalmology (Thyroid-associated ophthalmopathy) | 2023-2026 | 90 | NR | | | NR | NR | Low |
|  | D. Wang et al. (2026), China | Bilibili, Douyin | | Ophthalmology (Glaucoma) | 2023-2026 | 200 | NR | | | 355.67 | NR | Low |
|  | B. Kesimal et al. (2025), Turkey | Youtube | | Ophthalmology (Optic neuritis) | 2023-2026 | 50 | 60 | | | 161.6 | DISCERN, GQS, JAMA | Low |
|  | M. Kayabaşı et al. (2024), Turkey | Youtube | | Ophthalmology (Myopia) | 2023-2026 | 112 | 21 | | | 3174.84 | DISCERN, GQS, JAMA | Low |
|  | A. K. Sakallioglu et al. (2021), Turkey | Youtube | | Ophthalmology (Dry eye disease) | 2023-2026 | 238 | 23 | | | 611 | DISCERN, GQS | Low |

*Note: Data for engagement metrics are presented as Mean. Risk of bias was assessed using the Joanna Briggs Institute (JBI) Critical Appraisal Checklist for Analytical Cross-Sectional Studies. Studies were classified as Low, Moderate, or Low risk based on the proportion of met criteria. Search Period indicates the time frame during which the authors of the included studies retrieved the cross-sectional video data from the platforms. Abbreviations: NR, not reported.*
