## Supplementary material for "Quality of Chronic Disease–Related Health Videos Across Social Media Platforms: A Systematic Review and Meta-analysis": Table 2 Overall video quality analysis results

| **Subgroup Category** | **Studies (*k*)** | **Total Videos (*N*)** | **Pooled Score (95% CI)** | **Heterogeneity (I2)** | **Heterogeneity(τ2)** | **Heterogeneity (P-value)** |
| --- | --- | --- | --- | --- | --- | --- |
| Discern | 56 | 10767 | 50.61[48.04, 53.18] | 99.3 | 93.40 | *< 0.01* |
| GQS | 48 | 10680 | 56.93[54.44, 59.43] | 98.2 | 74.33 | *< 0.01* |
| JAMA | 25 | 6826 | 51.85[47.19, 56.52] | 98.7 | 138.21 | *< 0.01* |

*Note.* CI = Confidence Interval. Pooled scores were estimated using a random-effects model (REML) and represent the modified Percentage of Maximum Possible (POMP) score (standardized to a 0–100 scale).
