## Supplementary material for "Quality of Chronic Disease–Related Health Videos Across Social Media Platforms: A Systematic Review and Meta-analysis": Table 3 Subgroup Analysis of DISCERN by Platform Region

**Table 3. Subgroup Analysis of Standardized DISCERN Scores by Platform Region.**

| **Subgroup Category** | **Studies (*k*)** | **Total Videos (*N*)** | **Pooled Score (95% CI)** | **Heterogeneity (I2)** | **Heterogeneity(τ2)** | **Heterogeneity (P-value)** |
| --- | --- | --- | --- | --- | --- | --- |
| **Chinese Platform** | 35 | 8152 | 48.21[45.07, 51.36] | 99.4 | 88.36 | *< 0.01* |
| Bilibili | 32 | 2795 | 45.89[41.72, 50.07] | 99.3 | 140.67 | *< 0.01* |
| Douyin | 44 | 4917 | 49.85[46.66, 53.03] | 99.6 | 113.58 | *< 0.01* |
| Rednote | 8 | 1188 | 47.18[40.32, 54.04] | 98.3 | 96.02 | *< 0.01* |
| Kwai | 11 | 1830 | 47.36[41.09, 53.64] | 98.6 | 110.92 | *< 0.01* |
| **World Platform** | 21 | 2615 | 54.68[50.72, 58.64] | 98.7 | 81.19 | *< 0.01* |
| Youtube | 18 | 2300 | 56.91[53.42, 60.40] | 97.3 | 52.73 | *< 0.01* |
| Tiktok | 3 | 315 | 41.22[33.42, 49.01] | 95.3 | 45.57 | *< 0.01* |

*Note.* CI = Confidence Interval. Pooled scores were estimated using a random-effects model (REML) and represent the modified Percentage of Maximum Possible (POMP) score (standardized to a 0–100 scale). The test for subgroup differences showed a statistically significant difference between the two regions (P = 0.008). Because some included studies evaluated multiple platforms concurrently, the sum of *k* and *N* across specific sub-platforms may exceed the totals in the merged categories. The exceptionally high I^2^ across all subgroups indicates that traditional classifications (e.g., disease type) fail to fully explain the variance in video quality, highlighting the complex and highly volatile nature of the digital health communication ecosystem.
