## Supplementary material for "Quality of Chronic Disease–Related Health Videos Across Social Media Platforms: A Systematic Review and Meta-analysis": Table 4 Subgroup Analysis of DISCERN by Disease System

**Table 4. Subgroup Analysis of Standardized DISCERN Scores by Disease System.**

| **Disease System (Clinical Domain)** | **No. of Studies (Chs / Int)** | **Chinese Platforms (Mean, 95% CI)** | **International Platforms (Mean, 95% CI)** | **Test for Subgroup Differences (*p*-value)** |
| --- | --- | --- | --- | --- |
| Oncology | 9/4 | 50.06 [43.37, 56.76] | 58.01 [51.96, 64.05] | 0.08 |
| Musculoskeletal | 4/5 | 44.59 [37.50, 51.69] | 51.42 [42.06, 60.80] | 0.25 |
| Ophthalmology | 6/3 | 49.94 [43.51, 56.37] | 49.55 [44.36, 54.74] | 0.92 |

*Note:* Data are presented as Mean [95% CI]. Subgroups with fewer than 3 included studies (e.g., Digestive and Renal/Urinary systems in certain platforms) were excluded from the primary subgroup difference tests due to insufficient statistical power. The pooled estimates for these small subgroups are provided for descriptive purposes only and should not be interpreted as robust meta-analytical findings, as they are highly susceptible to single-study dominance and spurious precision.
